# Real-world evidence with a retrospective cohort of 15,968 Andalusian COVID-19 hospitalized patients suggests 21 new effective treatments and one drug that increases death risk

**DOI:** 10.1101/2022.08.14.22278751

**Authors:** Carlos Loucera, Rosario Carmona, Marina Esteban-Medina, Gerrit Bostelmann, Dolores Muñoyerro-Muñiz, Román Villegas, María Peña-Chilet, Joaquin Dopazo

## Abstract

Despite the extensive vaccination campaigns in many countries, COVID-19 is still a major worldwide health problem because of its associated morbidity and mortality. Therefore, finding efficient treatments as fast as possible is a pressing need. Drug repurposing constitutes a convenient alternative when the need for new drugs in an unexpected medical scenario is urgent, as is the case with COVID-19. Using data from a central registry of electronic health records (the Andalusian Population Health Database, BPS), the effect of prior consumption of drugs for other indications previous to the hospitalization with respect to patient survival was studied on a retrospective cohort of 15,968 individuals, comprising all COVID-19 patients hospitalized in Andalusia between January and November 2020. Covariate-adjusted hazard ratios and analysis of lymphocyte progression curves support a significant association between consumption of 21 different drugs and better patient survival. Contrarily, one drug, furosemide, displayed a significant increase in patient mortality.

## Introduction

During the COVID-19 pandemic, population-wide person-level electronic health record (EHR) data has increasingly gained importance for exploring, modeling, and reporting disease trends to inform healthcare and public health policy ^1^. The increasing availability of COVID-19 digital health data has fostered the interest in the use of real-world data (RWD) ^2^, defined as patient data collected from their EHRs, which can be analyzed to generate real-world evidence (RWE) ^3^. Actually, RWE can provide a better image of the actual clinical environments in which medical interventions are carried out when compared to conventional randomized clinical trials (RCTs), given that RWD includes detailed data on patient demographics, comorbidities, adherence, and simultaneous prescriptions ^4,5^. Moreover, RWE studies are not only cheaper than RCTs but can also be accomplished much faster, an advantage in scenarios in which an urgent decision must be taken, as in the case of a pandemic. In particular, discovering new drugs that could be used as efficient COVID-19 therapies is still an urgent need. Interestingly, much information on drugs, prescribed in COVID-19 patients for other indications, that could affect the progression of the disease is currently available in EHRs. For example, RWE has recently demonstrated that vitamin D has a significant protective effect on COVID-19 hospitalized patients ^6^. Therefore, RWD opens the door to carry out massive drug repurposing studies as well as research on potential adverse effects or interactions of drugs with COVID-19 progression.

Since 2001, the Andalusian Public Health System has systematically stored all the electronic health record (EHR) data of Andalusian patients in the Health Population Base (BPS) ^7^, which is currently one of the largest repositories of clinical data in the world (with over 13 million of comprehensive patient registries) ^7^. Because of its size and the detail of the data stored, BPS constitutes a unique and privileged environment to carry out large-scale RWE studies.

## Results and discussion

Clinical data for a total of 15,968 COVID-19 patients hospitalized in Andalusia between January and November 2020 were requested from the BPS. The data was transferred from BPS to the Infrastructure for secure real-world data analysis (iRWD)^8^ at the Foundation Progress and Health of the Andalusian Public Health System.

The endpoint considered was COVID-19 death during the first 30 days of hospital stay (see Methods). To elucidate if any given treatment could potentially reduce the mortality in COVID-19 inpatients a covariate balance analysis, which considers confounders (covariates that present an *a priori* possibility of confounding the association between a treatment and the survival outcome: sex, obesity, hypertension, cancer, pulmonary diseases, hypertension, asthma, age, and mental diseases; see Methods and Supplementary Table 1), was carried out to determinate the viability of further covariate-adjusted analysis. For these drugs eligible for covariate-adjusted analysis, survival was estimated using a weighted Cox Proportional Hazard model (See Methods), conditioned to the confounders of interest (Supplementary Table 1). A total of 864 treatments were identified in the BPS drug archive among the patients analyzed.

Since clinical data on laboratory analyses is also available in the BPS, lymphocyte progression, high levels of which account for a favorable progression, was assessed along with the drug treatment by a Linear Mixed Effects analysis, weighting the model with the same schema as in the survival analysis (see Methods for details).

Survival estimations showed that a total of 21 drugs have a significant effect on patient survival and, simultaneously, showed a significant increase in lymphocyte counts, after correction for the possible confounding covariables and for multiple testing (see Figure 1). Figure 2 shows the pattern of lymphocyte counts along the infection in the period studied for Enoxaparin (Figure 2A), which displays a clear trend of high levels of lymphocyte progression, for calcifediol (Figure 2B), with protective effect already reported ^6^, supported also by high levels of lymphocyte progression, and, as a counterexample, Furosemide, here linked to an increase in death risk, with lymphocyte levels below the average population.

**Figure 1.**
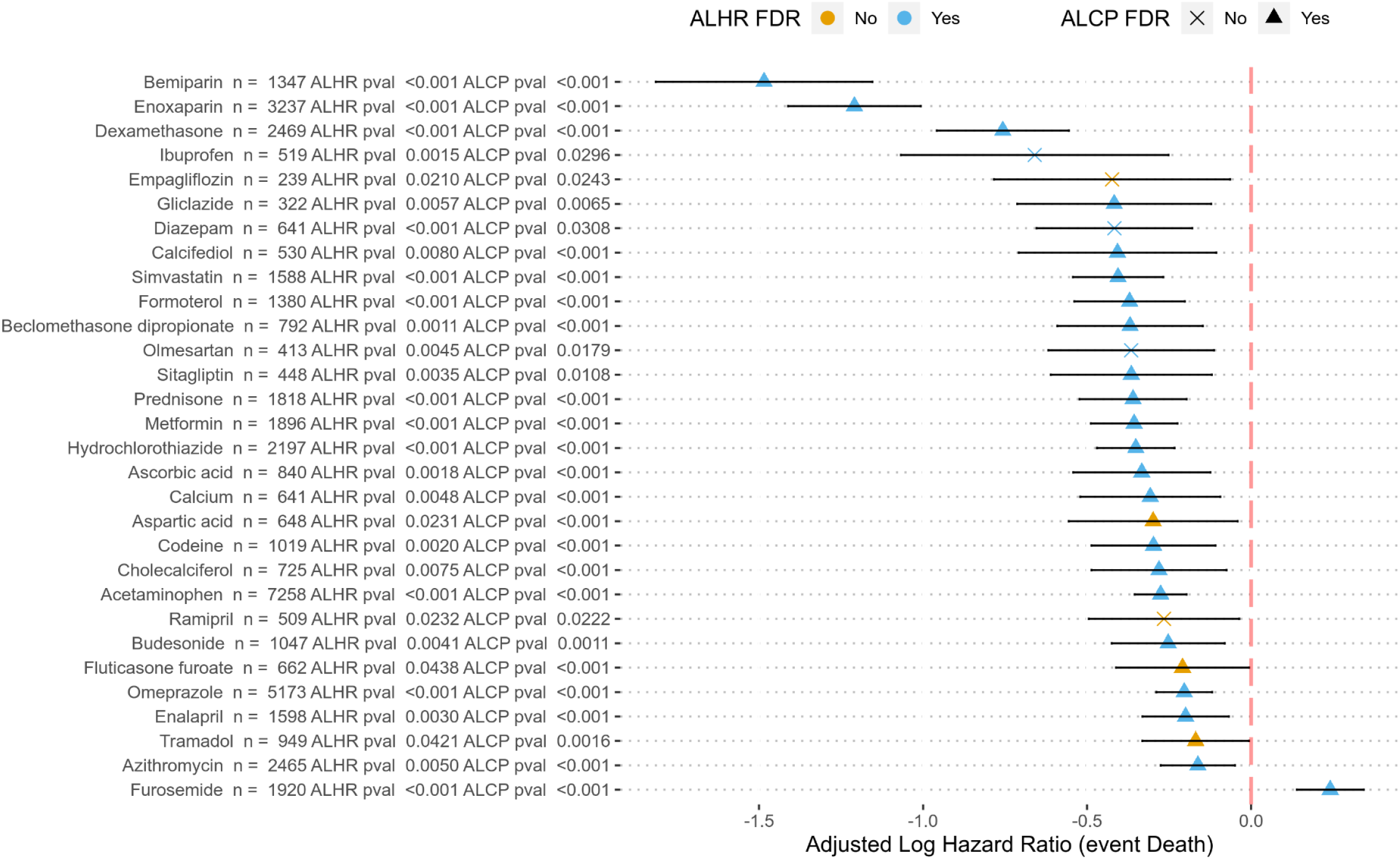
Adjusted log-hazard ratios 95% confidence intervals for all the eligible treatments that were significant in both analyses (survival and lymphocyte count progression) before and after FDR adjustment

**Figure 2.**
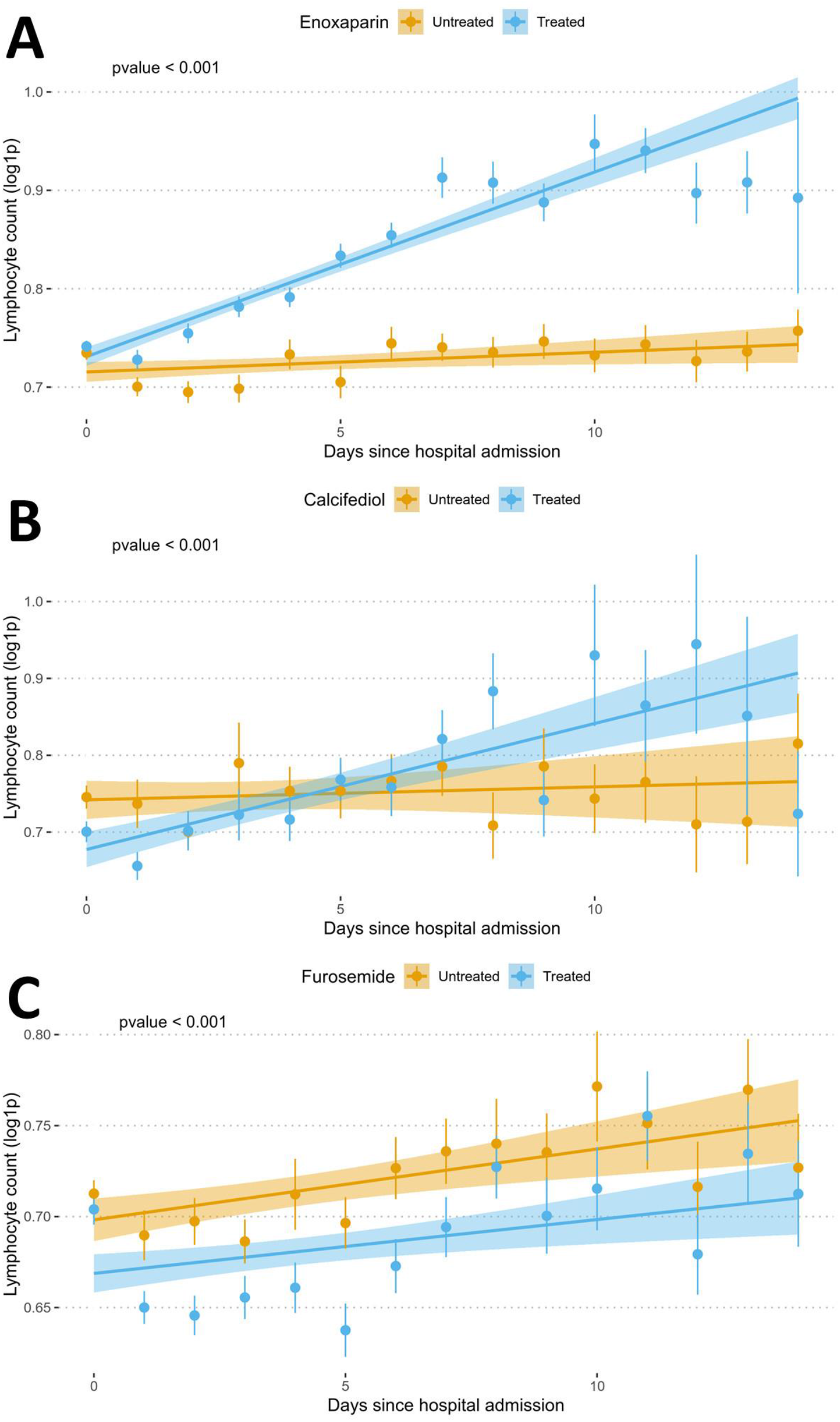
Plots showing the evolution of lymphocyte counts along the time studied (15 days since hospital admission) for A) enoxaparin, B) Calcifediol and C) Furosemide

The drugs associated to the highest survival, bemiparin (DB09258), LHR -1.62, 95% CI [-1.95,-1.31], FDR p-value=<10^−11^ and Enoxaparin (LHR -1.17, 95% CI [-1.36,-0.98], FDR p-value=<10^−11^), are antithrombotic used, as other heparins, to prevent thrombotic and thromboembolic complications in hospitalized patients. While for bemiparin no evidence of its protective effect has been found in the literature, a lower rate mortality in COVID-19 patients was described for enoxaparin when compared to other heparins^9^, in agreement with the results found here. However, this protective effect is not shared by other anticoagulants, such as Tinziparin (LHR= -0.34, 95% CI[-1.38, 0.69], FDR p-value= 1), despite its use in pulmonary embolism, or Fondaparinux (LHR=-0.33, 95% CI[-1.64, 0.97], FDR p-value=1). Calcifediol and Cholecalciferol, already described by us in a previous work ^6^, are significantly associated with better patient survival, probably due to the protective role of vitamin D due to its pro-immune and anti-inflammatory properties. Moreover, a number of the drugs found to affect COVID-19 patient survival were predicted as potentially active against COVID-19 ^10^ using machine learning and mathematical modeling ^11^ of the recently proposed COVID-19 the disease map^12^ (see the last two columns from Supplementary Table 2). It is interesting to note that, among the drugs eligible for the covariate-adjusted analysis (those in Supplementary Table 2) there is a significant enrichment of drugs predicted as repurposable by the machine learning model among those with a significant protective effect (X^2^ = 6.674, p-value = 0.009785), which supports the validity of the predictions previously made ^10^. One of drugs with a significant protective effect is simvastatin, a widely used statin, a group of drugs that reduce the blood level of low-density lipoprotein (LDL) cholesterol. Statins are also known for their pleiotropic effect, exerting an anti-inflammatory and antithrombotic action by inhibiting the NF-Kβ pathway which directly reduces inflammatory cytokines (IL1, IL6, TNF-α), CRP, and neutrophils^13^. Furthermore, a retrospective study performed in COVID-19 hospitalized patients showed that statins inhibit RAS activation and reduce angiotensin II proinflammatory effects, therefore improving endothelial function and remodeling after vascular injury ^14^. A recent *in-vitro* study demonstrates that simvastatin pretreatment in human Calu-3 epithelial lung cells inhibited SARS-CoV-2 binding and entry to the cell by inducing a redistribution of ACE2 receptors, lowering its concentration on the plasma membrane ^15^. Recent retrospective studies also point to the relationship between statin consumption and a reduced risk of mortality in COVID-19 patients ^14,16^. Another predicted drug is hydrochlorothiazide, a diuretic drug, often combined with ACE-inhibitors such as enalapril as antihypertensive therapy ^17^. It has been reported that patients with hypertension present a higher susceptibility to a severe COVID-19 prognosis ^18^, underlying hypertension as a risk factor for increased mortality in infected patients. Although the effect of antihypertensive drugs on COVID-19 patients with hypertension is controversial, the upregulation of ACE2 by ACE-inhibitors was linked to a dampened hyperinflammation and increased intrinsic antiviral responses of the cell in hypertensive COVID-19 patients ^19^. The results presented here, together with these previous reports, suggest that ACE-inhibitors may have a protective effect, in addition to contribute to a better survival in hypertensive patients.

## Conclusions

Although an exhaustive discussion of all the results found is beyond the scope of this report, the evidence provided by the covariate-adjusted analysis, reinforced by the lymphocyte analytics, strongly supports the findings presented here, based on RWD from a large retrospective cohort of 15,968 hospitalized patients across Andalusia. With a population of 8.5 million inhabitants, Andalusia is the third largest region in Europe and has a size comparable to countries like Switzerland or Austria. This population makes BPS, the database for secondary use of clinical data of the Andalusian Public Health System, a unique resource for large-scale RWE studies.

## Methods

### Design and patient selection

This study uses a retrospective cohort which includes Andalusian patients with COVID-19 diagnosis, hospitalized during the period January 2020 to November 2021.

The Ethics Committee for the Coordination of Biomedical Research in Andalusia approved the study “Retrospective analysis of all COVID-19 patients in the entire Andalusian community and generation of a prognostic predictor that can be applied preventively in possible future outbreaks” (29^th^ September, 2020, Acta 09/20) and the CEI from the University Hospitals Virgen Macarena and Virgen del Rocío approved the study “Medicina de precision en COVID-19 (PreMed-Covid19)” (22^nd^ December 2020, acta CEI 21/2020), and waived informed consent for the secondary use of clinical data for research purposes in both cases.

### Data management

Clinical data corresponding to COVID-19 patients hospitalized in Andalusia between January and November 2020 was requested to the Health Population Base (BPS), and from there transferred to the Infrastructure for secure real-world data analysis (iRWD) at the Foundation Progress and Health (FPS) of the Andalusian Public Health System for further analysis. In particular, the data listed in Supplementary Table 1 was extracted in BPS from the electronical health records for each patient and transferred to FPS for a total of 15,968 COVID-19 patients that fulfilled the inclusion criteria.

### Data preprocessing

Medication data in the office and hospital pharmacy records were found for 864 treatments. Individuals are considered as treated with a specific drug if prescriptions and the corresponding pharmacy dispensations (thereinafter a valid pharmacy order) were found within a period from 15 days before the hospital admission until the discharge up to 14 days (or death). Otherwise they were considered untreated.

The endpoint studied was COVID-19 death (certified death events during hospitalization). As in previous studies, the first 30 days of hospital stay were considered for survival calculations ^20^. The time variable in the models corresponds to the length (in days) of hospital stay. The stays that imply one or more changes of hospital units are combined in a single stay where the admission and discharge dates are set to either the start of the first or the end of the last combined stay. Only the first stay for each patient was considered to reduce potential biases due to reinfection.

### Covariate definition

Following previous studies^21^ the ICD codes were grouped into conditions as diabetes mellitus (ICD code E11), diseases of the circulatory system (ICD10 codes I00-I99), diseases of the respiratory system (ICD10 codes J00-J99), neoplasms (ICD10 codes C00-D49), dementia (ICD10 codes F00-F03), anxiety or mood disorders (ICD10 codes F30-F48), and other mental diseases (ICD10 codes F04-F29 and F50-F99). Obesity and other associated conditions (ICD10 codes D5-D8) with a possible confounding effect with the COVID-19 outcome were checked but no evidence was found in our database (nonsignificant χ^2^ association test). The age was categorized in the following ranks: [18, 40], [41, 67] and [68, 99). Gender was also considered as a known covariate. Table 1 displays the association between each covariate and the end point considered here, death, using chi-squared tests, along with the test p-value. Counts and proportions with respect to the end point are also provided.

**Table 1.** Association between each covariate and the end point using chi-squared tests, along with the test p-value, counts and proportions with respect to the end point.

| covariate | survival | death | p-value |
| --- | --- | --- | --- |
| Total N | 13116 | 2678 |  |
| Sex (female) | 6024 (45.9) | 1129 (42.2) | <0.001 |
| Flu vaccine | 5465 (41.7) | 1746 (65.2) | <0.001 |
| Pneumococcal vaccine | 3441 (26.2) | 1111 (41.5) | <0.001 |
| Diabetes | 3856 (29.4) | 1167 (43.6) | <0.001 |
| Circulatory diseases | 8111 (61.8) | 2261 (84.4) | <0.001 |
| Cancer | 1550 (11.8) | 545 (20.4) | <0.001 |
| Respiratory diseases | 2896 (22.1) | 828 (30.9) | <0.001 |
| Dementia | 964 (7.3) | 536 (20.0) | <0.001 |
| Other mental diseases | 1764 (13.4) | 407 (15.2) | 0.018 |
| Anxiety and mood disorders | 3382 (25.8) | 784 (29.3) | <0.001 |
| Age |  |  | <0.001 |
| 18_41 | 1399 (10.7) | 20 (0.7) |  |
| 41_68 | 5971 (45.5) | 380 (14.2) |  |
| 68_99 | 5746 (43.8) | 2278 (85.1) |  |

### Statistical analysis

To elucidate if any given treatment could potentially reduce the mortality in COVID-19 inpatients three statistical tests were conducted, considering covariates that present an a priori possibility of confounding the association between a treatment and the survival outcome ^22^ (see previous section).

Firstly, the survival outcome was estimated using a Cox Proportional Hazard model weighted using the inverse probability of treatment weighting (IPTW) technique, with the weights computed using a logistic regression model and adjusted for estimating the average treatment effect on the treated population (ATT) conditioned to the confounders of interest using the whole cohort. ATT is the most used weighting approximation to estimate treatment effects ^23^. To obtain an accurate measure of the variability of the marginal hazard ratios the closed-form estimator previously proposed ^24^ was used.

Then, the lymphocyte progression, a marker of COVID severity ^25^, is established as the different measurements of lymphocyte counts with respect to the initial day of hospitalization up to 14 days ^26^. Dates outside the hospitalization date range were omitted. The association between the positive linear trends in daily lymphocyte counts and reduced mortality in COVID-19 is obtained by comparing the trends in a treated population versus a control untreated population. A Linear Mixed Effects (LME) analysis was conducted to estimate if there was an increasing linear trend in the log-transformed lymphocyte progression due to being under a given treatment and the statistical significance was checked using an ANOVA analysis of the model ^27^. The model was weighted following the same weighting schema as in the survival analysis. In addition, a covariate balance analysis to determine the viability of the weighting schema ^28^ was carried out.

For each treatment, the inverse probability weighting (IPW) was used, based on propensity scores (IPW) generated using the WeightIt R package (v 0.12) ^29^. Here, the exposed condition is either having valid pharmacy order for the treatment during the 15 days prior to the beginning of the hospitalization event or during the first 14 days of the hospitalization. To assess the viability of the IPW analysis the proportion of covariates that could be effectively balanced was checked using the standardized mean differences test as implemented in the Cobalt R package (v 4.3.1) ^30^, using the 0.05 threshold ^28^. A treatment is eligible if all the covariates could be properly balanced, resulting in 122 eligible treatments out of the 864 initially found.

In both cases, p-values are corrected for multiple testing with False Discovery Rate (FDR) ^31^. Significance is achieved at level 0.05 and 95% confidence intervals are provided.

### Software

Weights for IPW are computed with the WeightIt R package (v 0.12) ^29^. IPW covariate suitability was computed using the Cobalt R package (v 4.3.1) ^30^. The survival estimation was conducted with R package HrIPW (v 0.1.2) ^32^. The LME analysis was conducted with R package lme4 (v 1.1-27) ^33^. The ANOVA analysis of the LME model was conducted with R package lmerTest (v 3.1-3) ^27^. R version 3.6.3 (2020-02-29).

## Data Availability

No data were generated in this study

## Acknowledgements

This research was funded by Spanish Ministry of Science and Innovation (grant PID2020-117979RB-I00), the Instituto de Salud Carlos III (ISCIII), co-funded with European Regional Development Funds (ERDF) (grant IMP/00019), and has also been funded by Consejería de Salud y Familias, Junta de Andalucía (grants COVID-0012-2020, PS-2020-342 and IE19_259 FPS) and the postdoctoral contract of Carlos Loucera (PAIDI2020-DOC_00350) co-funded by the European Social Fund (FSE) 2014-2020.

## Data Availability

The data supporting the findings of this study stored in the Andalusian Population Health Database (https://www.sspa.juntadeandalucia.es/servicioandaluzdesalud/profesionales/sistemas-de-informacion/base-poblacional-de-salud) but restrictions apply to the availability of these data, which were used under the ethics committee permission for the current study, and so are not publicly available.

## Supplementary Material

**Supplementary Table 1**. Data imported from BPS for each patient: code and definition of the variable.

**Supplementary Table 2**. Log Hazard ratios obtained for the drugs tested, along with standard deviations (SDs), upper and lower coefficient intervals (CI), nominal and FDR-adjusted p-values. Also, Lymphocyte proliferation values (see Methods) along with standard deviations (SDs), upper and lower coefficient intervals (CI), nominal and FDR-adjusted p-values. The two last columns indicate the drugs used in the machine learning drug repurposing prediction study ^10^ and the significance of the prediction.

