## Supplementary Table 1 for "Real-world evidence with a retrospective cohort of 15,968 Andalusian COVID-19 hospitalized patients suggests 21 new effective treatments and one drug that increases death risk"

**Supplementary Table 1**. Data imported from BPS for each patient: code and definition of the variable.

| Code | Meaning |
|---|---|
| FECNAC | Birth date |
| FECDEF | Death date |
| SEXO | Gender |
| FEC_INGRESO | Hospital admission date |
| FEC_ALTA | Discharge date |
| MOTIVO_ALTA | Reason for the discharge: (recovery/death/admission in another hospital/voluntary discharge/retirement home/unspecified) |
| COD_PATOLOGIA_CRONICA | Hospital codes for chronic conditions |
| COD_FEC_INI_PATOLOGIA | Date of condition diagnosis |
| COD_CIE_NORMALIZADO | A mixture of ICD9 and ICD10 codes for diseases |
| DESC_CIE_NORMALIZADO | Description of the ICD |
| FECINI_DIAG | Diagnosis date |
| FECFIN_DIAG | End of the diagnosed condition |
| FUENTE_DIAG | Source of the diagnosis (hospital, emergency, etc.) |
| IND_CRONICO_HCUP | Is a chronic disease? (yes/no) |
| Test COVID: FECHA | Test COVID date |
| Test COVID: TYPE | PCR / antigens |
| Test COVID: RESULTADO_TEST | Result of the test (positive/negative) |
| Pharmacy (Hospital and external): DESCRIPCION | List of drugs used in hospital or purchased in the pharmacies |
| Pharmacy (Hospital and external): FECHA | Dispensing date |
| VACUNA | List of vaccines |
| VACUNAFECHA | Vaccination dates |
| COD_CLC | List of analytical tests |
| FEC_EXTRAE | Analytic test date |
| VALOR | Analytic test value |
