## Supplementary table 2 for "Real-world evidence with a retrospective cohort of 15,968 Andalusian COVID-19 hospitalized patients suggests 21 new effective treatments and one drug that increases death risk"

**Supplementary Table 2**. Log Hazard ratios obtained for the drugs tested, along with standard deviations (SDs), upper and lower coefficient intervals (CI), nominal and FDR-adjusted p-values. Also, Lymphocyte proliferation values (see Methods) along with standard deviations (SDs), upper and lower coefficient intervals (CI), nominal and FDR-adjusted p-values. The two last columns indicate the drugs used in the machine learning drug repurposing prediction study and the significance of the prediction.

| Drugbank ID | Name | N | LHR estimate | LHR SD | LHR CI 5% | LHR CI 95% | LHR p-value | LHR FDR p-value | Lymphocyte proliferation | Lymphocyte Prol. CI 5% | Lymphocyte Prol. CI 95% | Lymphocyte Prol. p-value | Lymphocyte Prol. FDR p-value | Target | ML Prediction tested | ML Prediction significant |
|---|---|---|---|---|---|---|---|---|---|---|---|---|---|---|---|---|
| DB01225 | Enoxaparin | 3237 | -1.2092 | 0.1032 | -1.4114 | -1.0070 | 0.00E+00 | 0.00E+00 | 11.3107 | 0.0123 | 0.0175 | 5.06E-29 | 6.18E-27 | | Yes | No |
| DB09258 | Bemiparin | 1347 | -1.4845 | 0.1686 | -1.8149 | -1.1540 | 0.00E+00 | 0.00E+00 | 6.1915 | 0.0099 | 0.0191 | 8.87E-10 | 2.70E-08 | | Yes | No |
| DB01234 | Dexamethasone | 2469 | -0.7568 | 0.1027 | -0.9581 | -0.5554 | 1.75E-13 | 7.11E-12 | 9.4279 | 0.0123 | 0.0188 | 1.23E-20 | 7.53E-19 | NR1I2 | Yes | Yes |
| DB00316 | Acetaminophen | 7258 | -0.2757 | 0.0403 | -0.3548 | -0.1967 | 8.25E-12 | 2.52E-10 | 6.5357 | 0.0036 | 0.0067 | 6.80E-11 | 2.77E-09 | PTGS2 | Yes | Yes |
| DB00999 | Hydrochlorothiazide | 2197 | -0.3515 | 0.0604 | -0.4698 | -0.2332 | 5.78E-09 | 1.41E-07 | 3.9805 | 0.0027 | 0.0080 | 0.0001 | 0.0007 | KCNMA1 | Yes | Yes |
| DB00641 | Simvastatin | 1588 | -0.4056 | 0.0703 | -0.5433 | -0.2679 | 7.79E-09 | 1.58E-07 | 5.1663 | 0.0052 | 0.0116 | 2.71E-07 | 5.51E-06 | ITGB2 | Yes | Yes |
| DB00641 | Simvastatin | 1588 | -0.4056 | 0.0703 | -0.5433 | -0.2679 | 7.79E-09 | 1.58E-07 | 5.1663 | 0.0052 | 0.0116 | 2.71E-07 | 5.51E-06 | ITGAL | Yes | Yes |
| DB00331 | Metformin | 1896 | -0.3565 | 0.0673 | -0.4884 | -0.2245 | 1.19E-07 | 2.08E-06 | 3.8874 | 0.0027 | 0.0083 | 0.0001 | 0.0007 | GPD1 | Yes | Yes |
| DB00338 | Omeprazole | 5173 | -0.2037 | 0.0435 | -0.2890 | -0.1185 | 2.81E-06 | 4.29E-05 | 3.9230 | 0.0019 | 0.0057 | 0.0001 | 0.0007 | | Yes | No |
| DB00695 | Furosemide | 1920 | 0.2413 | 0.0521 | 0.1392 | 0.3435 | 3.66E-06 | 4.96E-05 | -3.7433 | -0.0081 | -0.0025 | 0.0002 | 0.0010 | | Yes | No |
| DB00635 | Prednisone | 1818 | -0.3598 | 0.0833 | -0.5231 | -0.1965 | 1.57E-05 | 0.0002 | 5.4336 | 0.0064 | 0.0137 | 6.41E-08 | 1.56E-06 | | Yes | No |
| DB00983 | Formoterol | 1380 | -0.3704 | 0.0860 | -0.5389 | -0.2019 | 1.64E-05 | 0.0002 | 5.1358 | 0.0065 | 0.0145 | 3.25E-07 | 5.66E-06 | | Yes | No |
| DB01611 | Hydroxychloroquine | 320 | -1.1799 | 0.3163 | -1.7999 | -0.5599 | 0.0002 | 0.0019 | -0.2891 | -0.0096 | 0.0073 | 0.7729 | 0.8129 | TLR7 | Yes | Yes |
| DB00381 | Amlodipine | 1250 | -0.2709 | 0.0729 | -0.4138 | -0.1280 | 0.0002 | 0.0019 | 1.8484 | -0.0002 | 0.0067 | 0.0648 | 0.1444 | CACNA1E | Yes | Yes |
| DB00381 | Amlodipine | 1250 | -0.2709 | 0.0729 | -0.4138 | -0.1280 | 0.0002 | 0.0019 | 1.8484 | -0.0002 | 0.0067 | 0.0648 | 0.1444 | CACNG1 | Yes | Yes |
| DB00945 | Acetylsalicylic acid | 2127 | -0.1954 | 0.0555 | -0.3042 | -0.0867 | 0.0004 | 0.0037 | 1.4022 | -0.0008 | 0.0048 | 0.1610 | 0.3070 | MYC | Yes | Yes |
| DB00945 | Acetylsalicylic acid | 2127 | -0.1954 | 0.0555 | -0.3042 | -0.0867 | 0.0004 | 0.0037 | 1.4022 | -0.0008 | 0.0048 | 0.1610 | 0.3070 | NFKBIA | Yes | Yes |
| DB00945 | Acetylsalicylic acid | 2127 | -0.1954 | 0.0555 | -0.3042 | -0.0867 | 0.0004 | 0.0037 | 1.4022 | -0.0008 | 0.0048 | 0.1610 | 0.3070 | CASP1 | Yes | Yes |
| DB00945 | Acetylsalicylic acid | 2127 | -0.1954 | 0.0555 | -0.3042 | -0.0867 | 0.0004 | 0.0037 | 1.4022 | -0.0008 | 0.0048 | 0.1610 | 0.3070 | IKBKB | Yes | Yes |
| DB00945 | Acetylsalicylic acid | 2127 | -0.1954 | 0.0555 | -0.3042 | -0.0867 | 0.0004 | 0.0037 | 1.4022 | -0.0008 | 0.0048 | 0.1610 | 0.3070 | CCND1 | Yes | Yes |
| DB00945 | Acetylsalicylic acid | 2127 | -0.1954 | 0.0555 | -0.3042 | -0.0867 | 0.0004 | 0.0037 | 1.4022 | -0.0008 | 0.0048 | 0.1610 | 0.3070 | PTGS2 | Yes | Yes |
| DB00945 | Acetylsalicylic acid | 2127 | -0.1954 | 0.0555 | -0.3042 | -0.0867 | 0.0004 | 0.0037 | 1.4022 | -0.0008 | 0.0048 | 0.1610 | 0.3070 | RPS6KA3 | Yes | Yes |

| | | | | | | | | | | | | | | | | |
|---|---|---|---|---|---|---|---|---|---|---|---|---|---|---|---|---|
| DB00829 | Diazepam | 641 | -0.4167 | 0.1211 | -0.6541 | -0.1793 | 0.0006 | 0.0047 | 2.1643 | 0.0006 | 0.0114 | 0.0308 | 0.0753 | GABRA2 | Yes | Yes |
| DB00829 | Diazepam | 641 | -0.4167 | 0.1211 | -0.6541 | -0.1793 | 0.0006 | 0.0047 | 2.1643 | 0.0006 | 0.0114 | 0.0308 | 0.0753 | GABRA5 | Yes | Yes |
| DB00829 | Diazepam | 641 | -0.4167 | 0.1211 | -0.6541 | -0.1793 | 0.0006 | 0.0047 | 2.1643 | 0.0006 | 0.0114 | 0.0308 | 0.0753 | GABRD | Yes | Yes |
| DB00394 | Beclomethasone dipropionate | 792 | -0.3690 | 0.1132 | -0.5909 | -0.1472 | 0.0011 | 0.0085 | 4.1539 | 0.0059 | 0.0166 | 3.65E-05 | 0.0004 | | Yes | No |
| DB01050 | Ibuprofen | 519 | -0.6596 | 0.2081 | -1.0674 | -0.2518 | 0.0015 | 0.0109 | 2.1828 | 0.0008 | 0.0150 | 0.0296 | 0.0737 | BCL2 | Yes | Yes |
| DB01050 | Ibuprofen | 519 | -0.6596 | 0.2081 | -1.0674 | -0.2518 | 0.0015 | 0.0109 | 2.1828 | 0.0008 | 0.0150 | 0.0296 | 0.0737 | PPARG | Yes | Yes |
| DB01050 | Ibuprofen | 519 | -0.6596 | 0.2081 | -1.0674 | -0.2518 | 0.0015 | 0.0109 | 2.1828 | 0.0008 | 0.0150 | 0.0296 | 0.0737 | CFTR | Yes | Yes |
| DB01050 | Ibuprofen | 519 | -0.6596 | 0.2081 | -1.0674 | -0.2518 | 0.0015 | 0.0109 | 2.1828 | 0.0008 | 0.0150 | 0.0296 | 0.0737 | PTGS2 | Yes | Yes |
| DB00126 | Ascorbic acid | 840 | -0.3332 | 0.1067 | -0.5424 | -0.1240 | 0.0018 | 0.0122 | 3.6668 | 0.0041 | 0.0134 | 0.0003 | 0.0013 | TMLHE | Yes | Yes |
| DB00126 | Ascorbic acid | 840 | -0.3332 | 0.1067 | -0.5424 | -0.1240 | 0.0018 | 0.0122 | 3.6668 | 0.0041 | 0.0134 | 0.0003 | 0.0013 | P3H1 | Yes | Yes |
| DB00126 | Ascorbic acid | 840 | -0.3332 | 0.1067 | -0.5424 | -0.1240 | 0.0018 | 0.0122 | 3.6668 | 0.0041 | 0.0134 | 0.0003 | 0.0013 | P4HTM | Yes | Yes |
| DB00126 | Ascorbic acid | 840 | -0.3332 | 0.1067 | -0.5424 | -0.1240 | 0.0018 | 0.0122 | 3.6668 | 0.0041 | 0.0134 | 0.0003 | 0.0013 | PHYH | Yes | Yes |
| DB00341 | Cetirizine | 233 | -0.7056 | 0.2279 | -1.1522 | -0.2590 | 0.0020 | 0.0125 | 0.9906 | -0.0046 | 0.0141 | 0.3223 | 0.4520 | HRH1 | Yes | Yes |
| DB00318 | Codeine | 1019 | -0.2975 | 0.0965 | -0.4865 | -0.1084 | 0.0020 | 0.0125 | 3.9242 | 0.0044 | 0.0132 | 0.0001 | 0.0007 | | Yes | No |
| DB00584 | Enalapril | 1598 | -0.1996 | 0.0673 | -0.3314 | -0.0678 | 0.0030 | 0.0174 | 3.7605 | 0.0030 | 0.0094 | 0.0002 | 0.0010 | | Yes | No |
| DB01261 | Sitagliptin | 448 | -0.3651 | 0.1251 | -0.6104 | -0.1198 | 0.0035 | 0.0196 | 2.5602 | 0.0018 | 0.0135 | 0.0108 | 0.0355 | DPP4 | Yes | Yes |
| DB01222 | Budesonide | 1047 | -0.2526 | 0.0880 | -0.4251 | -0.0801 | 0.0041 | 0.0217 | 3.2800 | 0.0029 | 0.0115 | 0.0011 | 0.0046 | | Yes | No |
| DB00706 | Tamsulosin | 1080 | 0.1925 | 0.0677 | 0.0599 | 0.3251 | 0.0044 | 0.0221 | 0.9666 | -0.0019 | 0.0055 | 0.3340 | 0.4630 | | Yes | No |
| DB00275 | Olmesartan | 413 | -0.3656 | 0.1288 | -0.6180 | -0.1132 | 0.0045 | 0.0221 | 2.3790 | 0.0014 | 0.0142 | 0.0179 | 0.0500 | AGTR1 | Yes | Yes |
| DB01373 | Calcium | 641 | -0.3073 | 0.1089 | -0.5208 | -0.0937 | 0.0048 | 0.0225 | 3.7516 | 0.0048 | 0.0153 | 0.0002 | 0.0010 | S100B | Yes | Yes |
| DB01373 | Calcium | 641 | -0.3073 | 0.1089 | -0.5208 | -0.0937 | 0.0048 | 0.0225 | 3.7516 | 0.0048 | 0.0153 | 0.0002 | 0.0010 | ALPP | Yes | Yes |
| DB01373 | Calcium | 641 | -0.3073 | 0.1089 | -0.5208 | -0.0937 | 0.0048 | 0.0225 | 3.7516 | 0.0048 | 0.0153 | 0.0002 | 0.0010 | PDCD6 | Yes | Yes |
| DB01373 | Calcium | 641 | -0.3073 | 0.1089 | -0.5208 | -0.0937 | 0.0048 | 0.0225 | 3.7516 | 0.0048 | 0.0153 | 0.0002 | 0.0010 | PCDH19 | Yes | Yes |
| DB01373 | Calcium | 641 | -0.3073 | 0.1089 | -0.5208 | -0.0937 | 0.0048 | 0.0225 | 3.7516 | 0.0048 | 0.0153 | 0.0002 | 0.0010 | CAST | Yes | Yes |
| DB01373 | Calcium | 641 | -0.3073 | 0.1089 | -0.5208 | -0.0937 | 0.0048 | 0.0225 | 3.7516 | 0.0048 | 0.0153 | 0.0002 | 0.0010 | S100A8 | Yes | Yes |
| DB01373 | Calcium | 641 | -0.3073 | 0.1089 | -0.5208 | -0.0937 | 0.0048 | 0.0225 | 3.7516 | 0.0048 | 0.0153 | 0.0002 | 0.0010 | COMP | Yes | Yes |
| DB00207 | Azithromycin | 2465 | -0.1622 | 0.0578 | -0.2754 | -0.0489 | 0.0050 | 0.0227 | 4.3675 | 0.0034 | 0.0089 | 1.32E-05 | 0.0002 | | Yes | No |
| DB01120 | Gliclazide | 322 | -0.4172 | 0.1510 | -0.7131 | -0.1213 | 0.0057 | 0.0249 | 2.7404 | 0.0026 | 0.0155 | 0.0065 | 0.0226 | | Yes | No |

| | | | | | | | | | | | | | | | | |
|---|---|---|---|---|---|---|---|---|---|---|---|---|---|---|---|---|
| DB00186 | Lorazepam | 1057 | -0.2300 | 0.0850 | -0.3966 | -0.0633 | 0.0068 | 0.0288 | 1.1617 | -0.0018 | 0.0071 | 0.2457 | 0.3996 | | Yes | No |
| DB00169 | Cholecalciferol | 725 | -0.2807 | 0.1050 | -0.4865 | -0.0748 | 0.0075 | 0.0306 | 3.6695 | 0.0044 | 0.0144 | 0.0003 | 0.0013 | VDR | Yes | Yes |
| DB00146 | Calcifediol | 530 | -0.4072 | 0.1536 | -0.7083 | -0.1061 | 0.0080 | 0.0316 | 3.4011 | 0.0050 | 0.0187 | 0.0007 | 0.0033 | VDR | Yes | Yes |
| DB01039 | Fenofibrate | 244 | -0.5385 | 0.2052 | -0.9407 | -0.1364 | 0.0087 | 0.0331 | 1.4990 | -0.0020 | 0.0147 | 0.1351 | 0.2659 | PPARG | Yes | Yes |
| DB01039 | Fenofibrate | 244 | -0.5385 | 0.2052 | -0.9407 | -0.1364 | 0.0087 | 0.0331 | 1.4990 | -0.0020 | 0.0147 | 0.1351 | 0.2659 | NR1I2 | Yes | Yes |
| DB00503 | Ritonavir | 199 | -2.5672 | 1.0184 | -4.5631 | -0.5712 | 0.0117 | 0.0433 | 0.5442 | -0.0073 | 0.0130 | 0.5870 | 0.6953 | NR1I2 | Yes | Yes |
| DB01601 | Lopinavir | 185 | -2.4728 | 1.0151 | -4.4623 | -0.4833 | 0.0148 | 0.0533 | 1.0065 | -0.0051 | 0.0159 | 0.3157 | 0.4520 | | No | No |
| DB01029 | Irbesartan | 184 | -0.4971 | 0.2119 | -0.9125 | -0.0817 | 0.0190 | 0.0662 | 0.6811 | -0.0066 | 0.0136 | 0.4962 | 0.6372 | JUN | Yes | Yes |
| DB01029 | Irbesartan | 184 | -0.4971 | 0.2119 | -0.9125 | -0.0817 | 0.0190 | 0.0662 | 0.6811 | -0.0066 | 0.0136 | 0.4962 | 0.6372 | AGTR1 | Yes | Yes |
| DB09154 | Sodium citrate | 272 | -0.4184 | 0.1802 | -0.7716 | -0.0652 | 0.0202 | 0.0670 | 0.8802 | -0.0045 | 0.0119 | 0.3795 | 0.5145 | | No | No |
| DB09341 | Dextrose, unspecified form | 311 | -0.3800 | 0.1638 | -0.7010 | -0.0590 | 0.0203 | 0.0670 | 0.7544 | -0.0046 | 0.0103 | 0.4512 | 0.5967 | | No | No |
| DB09038 | Empagliflozin | 239 | -0.4238 | 0.1836 | -0.7835 | -0.0640 | 0.0210 | 0.0673 | 2.2683 | 0.0013 | 0.0171 | 0.0243 | 0.0618 | | Yes | No |
| DB14500 | Potassium | 352 | -0.3476 | 0.1524 | -0.6464 | -0.0488 | 0.0226 | 0.0691 | 0.2757 | -0.0060 | 0.0080 | 0.7830 | 0.8164 | | No | No |
| DB00128 | Aspartic acid | 648 | -0.2984 | 0.1314 | -0.5560 | -0.0409 | 0.0231 | 0.0691 | 3.6651 | 0.0054 | 0.0179 | 0.0003 | 0.0013 | ASNS | Yes | Yes |
| DB00178 | Ramipril | 509 | -0.2656 | 0.1170 | -0.4949 | -0.0362 | 0.0232 | 0.0691 | 2.2942 | 0.0010 | 0.0129 | 0.0222 | 0.0602 | | Yes | No |
| DB09153 | Sodium chloride | 381 | -0.2960 | 0.1385 | -0.5674 | -0.0247 | 0.0325 | 0.0944 | 0.6257 | -0.0048 | 0.0093 | 0.5319 | 0.6603 | | No | No |
| DB00193 | Tramadol | 949 | -0.1690 | 0.0831 | -0.3319 | -0.0060 | 0.0421 | 0.1192 | 3.1635 | 0.0025 | 0.0107 | 0.0016 | 0.0064 | SLC6A4 | Yes | Yes |
| DB08906 | Fluticasone furoate | 662 | -0.2089 | 0.1036 | -0.4120 | -0.0058 | 0.0438 | 0.1192 | 4.0096 | 0.0055 | 0.0159 | 0.0001 | 0.0007 | | Yes | No |
| DB01098 | Rosuvastatin | 314 | -0.3010 | 0.1494 | -0.5939 | -0.0081 | 0.0440 | 0.1192 | 1.0299 | -0.0038 | 0.0121 | 0.3039 | 0.4423 | | Yes | No |
| DB01418 | Acenocoumarol | 511 | 0.1784 | 0.0897 | 0.0025 | 0.3543 | 0.0468 | 0.1241 | -0.7458 | -0.0079 | 0.0035 | 0.4561 | 0.5967 | | Yes | No |
| DB13872 | Lormetazepam | 806 | 0.1523 | 0.0779 | -0.0003 | 0.3049 | 0.0504 | 0.1289 | 0.3720 | -0.0037 | 0.0054 | 0.7100 | 0.7666 | GABRA2 | Yes | Yes |
| DB13872 | Lormetazepam | 806 | 0.1523 | 0.0779 | -0.0003 | 0.3049 | 0.0504 | 0.1289 | 0.3720 | -0.0037 | 0.0054 | 0.7100 | 0.7666 | GABRA5 | Yes | Yes |
| DB00455 | Loratadine | 251 | -0.3619 | 0.1852 | -0.7249 | 0.0011 | 0.0507 | 0.1289 | 1.3378 | -0.0029 | 0.0155 | 0.1817 | 0.3309 | HRH1 | Yes | Yes |
| DB00654 | Latanoprost | 183 | 0.2849 | 0.1475 | -0.0042 | 0.5740 | 0.0534 | 0.1329 | -0.9337 | -0.0143 | 0.0048 | 0.3521 | 0.4827 | | Yes | No |
| DB00537 | Ciprofloxacin | 353 | 0.2213 | 0.1151 | -0.0042 | 0.4468 | 0.0545 | 0.1329 | 1.0522 | -0.0035 | 0.0117 | 0.2934 | 0.4419 | | Yes | No |
| DB01126 | Dutasteride | 547 | 0.1649 | 0.0881 | -0.0078 | 0.3377 | 0.0613 | 0.1467 | 1.4243 | -0.0014 | 0.0088 | 0.1549 | 0.3000 | SRD5A1 | Yes | Yes |
| DB00656 | Trazodone | 468 | 0.1784 | 0.0965 | -0.0109 | 0.3676 | 0.0647 | 0.1499 | 0.2636 | -0.0053 | 0.0070 | 0.7922 | 0.8191 | HRH1 | Yes | Yes |
| DB00656 | Trazodone | 468 | 0.1784 | 0.0965 | -0.0109 | 0.3676 | 0.0647 | 0.1499 | 0.2636 | -0.0053 | 0.0070 | 0.7922 | 0.8191 | SLC6A4 | Yes | Yes |

| | | | | | | | | | | | | | | | | |
|---|---|---|---|---|---|---|---|---|---|---|---|---|---|---|---|---|
| DB01306 | Insulin aspart | 214 | -0.3385 | 0.1835 | -0.6981 | 0.0212 | 0.0651 | 0.1499 | 2.0462 | 0.0004 | 0.0186 | 0.0420 | 0.1004 | | Yes | No |
| DB06292 | Dapagliflozin | 229 | -0.3537 | 0.1933 | -0.7326 | 0.0251 | 0.0672 | 0.1519 | 2.5051 | 0.0024 | 0.0194 | 0.0131 | 0.0410 | | Yes | No |
| DB00727 | Nitroglycerin | 359 | 0.2059 | 0.1136 | -0.0168 | 0.4286 | 0.0700 | 0.1553 | -1.2818 | -0.0115 | 0.0024 | 0.2011 | 0.3555 | | Yes | No |
| DB00736 | Esomeprazole | 277 | -0.2659 | 0.1573 | -0.5743 | 0.0425 | 0.0911 | 0.1984 | 1.5575 | -0.0018 | 0.0158 | 0.1204 | 0.2408 | | Yes | No |
| DB00678 | Losartan | 1409 | -0.1090 | 0.0665 | -0.2393 | 0.0213 | 0.1011 | 0.2165 | 3.9044 | 0.0033 | 0.0099 | 0.0001 | 0.0007 | AGTR1 | Yes | Yes |
| DB00335 | Atenolol | 187 | -0.3154 | 0.1935 | -0.6946 | 0.0639 | 0.1031 | 0.2170 | -0.6043 | -0.0126 | 0.0067 | 0.5464 | 0.6666 | | Yes | No |
| DB00796 | Candesartan cilexetil | 198 | -0.3031 | 0.1952 | -0.6856 | 0.0794 | 0.1204 | 0.2489 | 2.5196 | 0.0031 | 0.0246 | 0.0126 | 0.0405 | AGTR1 | Yes | Yes |
| DB08882 | Linagliptin | 306 | 0.1743 | 0.1170 | -0.0550 | 0.4037 | 0.1362 | 0.2690 | -0.4302 | -0.0092 | 0.0059 | 0.6673 | 0.7609 | DPP4 | Yes | Yes |
| DB09214 | Dexketoprofen | 327 | -0.2547 | 0.1710 | -0.5899 | 0.0805 | 0.1364 | 0.2690 | 1.0284 | -0.0043 | 0.0137 | 0.3045 | 0.4423 | PTGS2 | Yes | Yes |
| DB00451 | Levothyroxine | 396 | -0.2245 | 0.1509 | -0.5202 | 0.0712 | 0.1367 | 0.2690 | 2.2650 | 0.0011 | 0.0152 | 0.0241 | 0.0618 | THRB | Yes | Yes |
| DB00373 | Timolol | 254 | -0.2166 | 0.1475 | -0.5057 | 0.0725 | 0.1421 | 0.2751 | -0.3720 | -0.0118 | 0.0081 | 0.7100 | 0.7666 | | Yes | No |
| DB00047 | Insulin glargine | 565 | -0.1516 | 0.1044 | -0.3562 | 0.0529 | 0.1463 | 0.2789 | 1.1004 | -0.0023 | 0.0082 | 0.2716 | 0.4195 | | Yes | No |
| DB00986 | Glycopyrronium | 196 | 0.2022 | 0.1423 | -0.0768 | 0.4812 | 0.1555 | 0.2919 | 0.4003 | -0.0078 | 0.0118 | 0.6894 | 0.7666 | | Yes | No |
| DB00788 | Naproxen | 223 | -0.2915 | 0.2101 | -0.7032 | 0.1202 | 0.1653 | 0.3055 | 3.2195 | 0.0059 | 0.0243 | 0.0015 | 0.0063 | PTGS2 | Yes | Yes |
| DB09082 | Vilanterol | 328 | -0.2052 | 0.1492 | -0.4975 | 0.0872 | 0.1690 | 0.3078 | 2.2803 | 0.0012 | 0.0165 | 0.0234 | 0.0618 | | Yes | No |
| DB00813 | Fentanyl | 251 | 0.1688 | 0.1263 | -0.0787 | 0.4163 | 0.1812 | 0.3252 | 0.1296 | -0.0083 | 0.0095 | 0.8970 | 0.9119 | | Yes | No |
| DB00590 | Doxazosin | 428 | -0.1451 | 0.1128 | -0.3662 | 0.0761 | 0.1985 | 0.3510 | 1.0916 | -0.0027 | 0.0095 | 0.2755 | 0.4202 | | Yes | No |
| DB04817 | Metamizole | 2843 | -0.0640 | 0.0507 | -0.1635 | 0.0355 | 0.2072 | 0.3612 | 4.0654 | 0.0027 | 0.0076 | 4.93E-05 | 0.0005 | | No | No |
| DB00973 | Ezetimibe | 344 | -0.1615 | 0.1308 | -0.4178 | 0.0947 | 0.2166 | 0.3723 | 1.8670 | -0.0004 | 0.0163 | 0.0626 | 0.1444 | SOAT1 | Yes | Yes |
| DB00448 | Lansoprazole | 236 | -0.1829 | 0.1510 | -0.4789 | 0.1131 | 0.2258 | 0.3825 | -0.3278 | -0.0101 | 0.0072 | 0.7434 | 0.7886 | | Yes | No |
| DB01175 | Escitalopram | 175 | 0.2005 | 0.1673 | -0.1273 | 0.5283 | 0.2307 | 0.3855 | 2.8917 | 0.0055 | 0.0289 | 0.0044 | 0.0161 | HRH1 | Yes | Yes |
| DB01175 | Escitalopram | 175 | 0.2005 | 0.1673 | -0.1273 | 0.5283 | 0.2307 | 0.3855 | 2.8917 | 0.0055 | 0.0289 | 0.0044 | 0.0161 | SLC6A4 | Yes | Yes |
| DB01076 | Atorvastatin | 1112 | -0.0845 | 0.0735 | -0.2286 | 0.0597 | 0.2507 | 0.4089 | 1.1407 | -0.0016 | 0.0062 | 0.2543 | 0.4029 | DPP4 | Yes | Yes |
| DB00927 | Famotidine | 207 | -0.1926 | 0.1679 | -0.5218 | 0.1365 | 0.2514 | 0.4089 | 0.5732 | -0.0072 | 0.0132 | 0.5670 | 0.6849 | | Yes | No |
| DB01001 | Salbutamol | 670 | -0.1196 | 0.1097 | -0.3346 | 0.0953 | 0.2753 | 0.4419 | 3.9062 | 0.0057 | 0.0171 | 0.0001 | 0.0007 | | Yes | No |
| DB00421 | Spironolactone | 272 | 0.1233 | 0.1237 | -0.1191 | 0.3657 | 0.3189 | 0.5023 | 0.4487 | -0.0051 | 0.0081 | 0.6540 | 0.7599 | NR1I2 | Yes | Yes |
| DB00421 | Spironolactone | 272 | 0.1233 | 0.1237 | -0.1191 | 0.3657 | 0.3189 | 0.5023 | 0.4487 | -0.0051 | 0.0081 | 0.6540 | 0.7599 | CACNG1 | Yes | Yes |
| DB01558 | Bromazepam | 471 | -0.1147 | 0.1156 | -0.3413 | 0.1119 | 0.3211 | 0.5023 | 2.7179 | 0.0023 | 0.0145 | 0.0068 | 0.0231 | GABRA2 | Yes | Yes |

| | | | | | | | | | | | | | | | | |
|---|---|---|---|---|---|---|---|---|---|---|---|---|---|---|---|---|
| DB01558 | Bromazepam | 471 | -0.1147 | 0.1156 | -0.3413 | 0.1119 | 0.3211 | 0.5023 | 2.7179 | 0.0023 | 0.0145 | 0.0068 | 0.0231 | GABRA5 | Yes | Yes |
| DB01558 | Bromazepam | 471 | -0.1147 | 0.1156 | -0.3413 | 0.1119 | 0.3211 | 0.5023 | 2.7179 | 0.0023 | 0.0145 | 0.0068 | 0.0231 | GABRA6 | Yes | Yes |
| DB01558 | Bromazepam | 471 | -0.1147 | 0.1156 | -0.3413 | 0.1119 | 0.3211 | 0.5023 | 2.7179 | 0.0023 | 0.0145 | 0.0068 | 0.0231 | GABRD | Yes | Yes |
| DB01592 | Iron | 959 | 0.0757 | 0.0774 | -0.0759 | 0.2274 | 0.3275 | 0.5058 | -1.3404 | -0.0079 | 0.0015 | 0.1805 | 0.3309 | FEN1 | Yes | Yes |
| DB00215 | Citalopram | 300 | -0.1365 | 0.1427 | -0.4162 | 0.1433 | 0.3391 | 0.5126 | 2.8528 | 0.0036 | 0.0197 | 0.0046 | 0.0167 | HRH1 | Yes | Yes |
| DB00215 | Citalopram | 300 | -0.1365 | 0.1427 | -0.4162 | 0.1433 | 0.3391 | 0.5126 | 2.8528 | 0.0036 | 0.0197 | 0.0046 | 0.0167 | SLC6A4 | Yes | Yes |
| DB00177 | Valsartan | 383 | -0.1236 | 0.1296 | -0.3777 | 0.1305 | 0.3403 | 0.5126 | 1.8581 | -0.0003 | 0.0126 | 0.0639 | 0.1444 | AGTR1 | Yes | Yes |
| DB01184 | Domperidone | 226 | 0.1325 | 0.1483 | -0.1581 | 0.4231 | 0.3715 | 0.5527 | 1.6460 | -0.0015 | 0.0174 | 0.1010 | 0.2088 | | Yes | No |
| DB00828 | Fosfomycin | 298 | -0.1253 | 0.1462 | -0.4118 | 0.1612 | 0.3913 | 0.5751 | 0.5452 | -0.0053 | 0.0094 | 0.5862 | 0.6953 | | No | No |
| DB04876 | Vildagliptin | 200 | -0.1319 | 0.1611 | -0.4476 | 0.1838 | 0.4129 | 0.5996 | 2.3836 | 0.0018 | 0.0181 | 0.0180 | 0.0500 | DPP4 | Yes | Yes |
| DB09238 | Manidipine | 160 | -0.1399 | 0.1886 | -0.5095 | 0.2297 | 0.4583 | 0.6578 | -0.3434 | -0.0119 | 0.0084 | 0.7317 | 0.7831 | | Yes | No |
| DB00437 | Allopurinol | 511 | -0.0686 | 0.1010 | -0.2666 | 0.1295 | 0.4974 | 0.7056 | -0.6197 | -0.0067 | 0.0035 | 0.5358 | 0.6603 | | Yes | No |
| DB11921 | Deflazacort | 629 | -0.0655 | 0.1022 | -0.2658 | 0.1348 | 0.5216 | 0.7250 | 3.8284 | 0.0052 | 0.0162 | 0.0001 | 0.0009 | | Yes | No |
| DB00612 | Bisoprolol | 1409 | 0.0395 | 0.0619 | -0.0817 | 0.1608 | 0.5230 | 0.7250 | 0.4544 | -0.0026 | 0.0042 | 0.6496 | 0.7599 | | Yes | No |
| DB00966 | Telmisartan | 226 | -0.0961 | 0.1565 | -0.4030 | 0.2107 | 0.5392 | 0.7363 | 1.2578 | -0.0031 | 0.0141 | 0.2099 | 0.3652 | PPARG | Yes | Yes |
| DB00966 | Telmisartan | 226 | -0.0961 | 0.1565 | -0.4030 | 0.2107 | 0.5392 | 0.7363 | 1.2578 | -0.0031 | 0.0141 | 0.2099 | 0.3652 | AGTR1 | Yes | Yes |
| DB00646 | Nystatin | 165 | -0.1252 | 0.2059 | -0.5287 | 0.2784 | 0.5432 | 0.7363 | -1.1110 | -0.0193 | 0.0054 | 0.2679 | 0.4190 | | No | No |
| DB06228 | Rivaroxaban | 200 | -0.0944 | 0.1629 | -0.4137 | 0.2250 | 0.5625 | 0.7541 | 0.7740 | -0.0058 | 0.0134 | 0.4399 | 0.5897 | | Yes | No |
| DB00332 | Ipratropium | 1322 | -0.0410 | 0.0748 | -0.1876 | 0.1056 | 0.5834 | 0.7736 | 5.0385 | 0.0063 | 0.0144 | 5.53E-07 | 8.44E-06 | | Yes | No |
| DB00214 | Torasemide | 278 | 0.0708 | 0.1323 | -0.1886 | 0.3302 | 0.5927 | 0.7775 | 1.2194 | -0.0030 | 0.0127 | 0.2238 | 0.3735 | | Yes | No |
| DB01137 | Levofloxacin | 1052 | 0.0380 | 0.0749 | -0.1089 | 0.1848 | 0.6124 | 0.7859 | 3.7623 | 0.0039 | 0.0124 | 0.0002 | 0.0010 | | Yes | No |
| DB00213 | Pantoprazole | 476 | 0.0516 | 0.1036 | -0.1515 | 0.2546 | 0.6187 | 0.7859 | 1.7043 | -0.0008 | 0.0113 | 0.0890 | 0.1873 | | Yes | No |
| DB00440 | Trimethoprim | 161 | -0.1046 | 0.2113 | -0.5187 | 0.3095 | 0.6206 | 0.7859 | 2.4377 | 0.0028 | 0.0254 | 0.0157 | 0.0480 | | Yes | No |
| DB00630 | Alendronic acid | 172 | -0.0888 | 0.1817 | -0.4450 | 0.2673 | 0.6248 | 0.7859 | 1.2955 | -0.0039 | 0.0192 | 0.1968 | 0.3532 | | Yes | No |
| DB01112 | Cefuroxime | 352 | 0.0628 | 0.1319 | -0.1956 | 0.3213 | 0.6338 | 0.7890 | 1.2115 | -0.0028 | 0.0119 | 0.2265 | 0.3735 | | No | No |
| DB01233 | Metoclopramide | 498 | 0.0565 | 0.1234 | -0.1854 | 0.2983 | 0.6473 | 0.7977 | 2.3802 | 0.0014 | 0.0150 | 0.0177 | 0.0500 | HTR4 | Yes | Yes |
| DB00758 | Clopidogrel | 285 | 0.0539 | 0.1283 | -0.1975 | 0.3053 | 0.6745 | 0.8155 | -0.3938 | -0.0083 | 0.0055 | 0.6940 | 0.7666 | | Yes | No |
| DB01068 | Clonazepam | 167 | 0.0751 | 0.1792 | -0.2761 | 0.4263 | 0.6751 | 0.8155 | -0.1190 | -0.0123 | 0.0109 | 0.9054 | 0.9129 | NR1I2 | Yes | Yes |

| | | | | | | | | | | | | | | | | |
|---|---|---|---|---|---|---|---|---|---|---|---|---|---|---|---|---|
| DB00766 | Clavulanic acid | 779 | 0.0367 | 0.0908 | -0.1413 | 0.2146 | 0.6864 | 0.8209 | 1.0387 | -0.0026 | 0.0083 | 0.2995 | 0.4423 | | No | No |
| DB00230 | Pregabalin | 382 | 0.0402 | 0.1138 | -0.1828 | 0.2632 | 0.7240 | 0.8575 | -0.3773 | -0.0077 | 0.0052 | 0.7061 | 0.7666 | | Yes | No |
| DB00370 | Mirtazapine | 201 | 0.0522 | 0.1522 | -0.2460 | 0.3504 | 0.7317 | 0.8583 | -0.3812 | -0.0104 | 0.0070 | 0.7035 | 0.7666 | HRH1 | Yes | Yes |
| DB00502 | Haloperidol | 206 | 0.0380 | 0.1436 | -0.2435 | 0.3194 | 0.7913 | 0.9113 | 1.8544 | -0.0004 | 0.0153 | 0.0651 | 0.1444 | | Yes | No |
| DB01136 | Carvedilol | 370 | -0.0309 | 0.1172 | -0.2605 | 0.1987 | 0.7918 | 0.9113 | -0.9917 | -0.0092 | 0.0030 | 0.3220 | 0.4520 | HIF1A | Yes | Yes |
| DB00628 | Clorazepic acid | 330 | -0.0323 | 0.1286 | -0.2843 | 0.2198 | 0.8018 | 0.9142 | 0.6315 | -0.0048 | 0.0093 | 0.5282 | 0.6603 | | Yes | No |
| DB08810 | Cinitapride | 210 | -0.0410 | 0.1783 | -0.3905 | 0.3084 | 0.8180 | 0.9240 | 2.4107 | 0.0022 | 0.0210 | 0.0168 | 0.0500 | | No | No |
| DB14761 | Remdesivir | 190 | -0.0396 | 0.2061 | -0.4435 | 0.3643 | 0.8477 | 0.9488 | 1.5787 | -0.0018 | 0.0161 | 0.1160 | 0.2359 | | No | No |
| DB01409 | Tiotropium | 449 | -0.0191 | 0.1091 | -0.2330 | 0.1947 | 0.8607 | 0.9546 | 0.4313 | -0.0048 | 0.0075 | 0.6665 | 0.7609 | | Yes | No |
| DB00715 | Paroxetine | 174 | -0.0290 | 0.1960 | -0.4131 | 0.3552 | 0.8825 | 0.9571 | 1.2141 | -0.0037 | 0.0160 | 0.2263 | 0.3735 | SLC6A4 | Yes | Yes |
| DB00158 | Folic acid | 624 | -0.0149 | 0.1022 | -0.2152 | 0.1855 | 0.8843 | 0.9571 | -0.6695 | -0.0080 | 0.0039 | 0.5035 | 0.6398 | FOLR1 | Yes | Yes |
| DB06605 | Apixaban | 345 | -0.0161 | 0.1184 | -0.2482 | 0.2160 | 0.8919 | 0.9571 | 0.7404 | -0.0043 | 0.0095 | 0.4598 | 0.5967 | | Yes | No |
| DB06698 | Betahistine | 409 | -0.0153 | 0.1154 | -0.2415 | 0.2109 | 0.8944 | 0.9571 | 1.3829 | -0.0019 | 0.0109 | 0.1675 | 0.3145 | HRH1 | Yes | Yes |
| DB00806 | Pentoxifylline | 231 | -0.0155 | 0.1376 | -0.2852 | 0.2542 | 0.9104 | 0.9658 | -0.2132 | -0.0088 | 0.0071 | 0.8314 | 0.8523 | NT5E | Yes | Yes |
| DB00425 | Zolpidem | 292 | -0.0095 | 0.1360 | -0.2760 | 0.2569 | 0.9441 | 0.9760 | 1.8193 | -0.0006 | 0.0146 | 0.0704 | 0.1533 | GABRA2 | Yes | Yes |
| DB01060 | Amoxicillin | 1090 | 0.0052 | 0.0779 | -0.1475 | 0.1578 | 0.9472 | 0.9760 | 1.2477 | -0.0015 | 0.0068 | 0.2125 | 0.3652 | | No | No |
| DB00404 | Alprazolam | 557 | -0.0058 | 0.1044 | -0.2105 | 0.1989 | 0.9558 | 0.9760 | 3.2861 | 0.0037 | 0.0145 | 0.0011 | 0.0046 | GABRA2 | Yes | Yes |
| DB00404 | Alprazolam | 557 | -0.0058 | 0.1044 | -0.2105 | 0.1989 | 0.9558 | 0.9760 | 3.2861 | 0.0037 | 0.0145 | 0.0011 | 0.0046 | GABRA5 | Yes | Yes |
| DB00404 | Alprazolam | 557 | -0.0058 | 0.1044 | -0.2105 | 0.1989 | 0.9558 | 0.9760 | 3.2861 | 0.0037 | 0.0145 | 0.0011 | 0.0046 | GABRA6 | Yes | Yes |
| DB00321 | Amitriptyline | 162 | 0.0088 | 0.1867 | -0.3571 | 0.3747 | 0.9623 | 0.9760 | 1.7201 | -0.0013 | 0.0194 | 0.0875 | 0.1872 | NTRK1 | Yes | Yes |
| DB00321 | Amitriptyline | 162 | 0.0088 | 0.1867 | -0.3571 | 0.3747 | 0.9623 | 0.9760 | 1.7201 | -0.0013 | 0.0194 | 0.0875 | 0.1872 | HTR1B | Yes | Yes |
| DB00321 | Amitriptyline | 162 | 0.0088 | 0.1867 | -0.3571 | 0.3747 | 0.9623 | 0.9760 | 1.7201 | -0.0013 | 0.0194 | 0.0875 | 0.1872 | HRH1 | Yes | Yes |
| DB00321 | Amitriptyline | 162 | 0.0088 | 0.1867 | -0.3571 | 0.3747 | 0.9623 | 0.9760 | 1.7201 | -0.0013 | 0.0194 | 0.0875 | 0.1872 | SLC6A4 | Yes | Yes |
| DB00115 | Cyanocobalamin | 400 | -0.0056 | 0.1412 | -0.2824 | 0.2712 | 0.9685 | 0.9760 | 0.0728 | -0.0084 | 0.0090 | 0.9420 | 0.9420 | | Yes | No |
| DB00938 | Salmeterol | 279 | 0.0048 | 0.1348 | -0.2594 | 0.2690 | 0.9715 | 0.9760 | 1.1509 | -0.0035 | 0.0135 | 0.2507 | 0.4025 | | Yes | No |
| DB01104 | Sertraline | 359 | -0.0037 | 0.1213 | -0.2415 | 0.2342 | 0.9760 | 0.9760 | 2.9841 | 0.0036 | 0.0175 | 0.0030 | 0.0116 | SLC6A4 | Yes | Yes |
